# Comparison between Propofol and Total Inhalational Anaesthesia on Cardiovascular Outcomes Following On-pump Cardiac Surgery in Higher-Risk Patients – A Randomised Controlled Pilot and Feasibility Study

**DOI:** 10.1101/2024.02.04.24302307

**Authors:** Benjamin Milne, Martin John, Richard Evans, Steven Robertson, Pádraig Ó Scanaill, Gavin J Murphy, Giovanni Landoni, Mike Marber, Tim Clayton, Gudrun Kunst

## Abstract

**Objectives:** Myocardial revascularisation and cardiopulmonary bypass (CPB) can cause ischemia-reperfusion injury, leading to myocardial and other end-organ damage. Volatile anaesthetics protect the myocardium in experimental studies, however there is uncertainty as whether this translates into clinical benefits.

**Methods:** In this single blinded parallel group randomised controlled feasibility trial higher-risk patients undergoing elective coronary artery bypass graft surgery with an additive European System for Cardiac Operative Risk Evaluation (EuroScore) ≥ 5 were randomised to receive either propofol or total inhalational anaesthesia for maintenance of anaesthesia. The primary outcome was the feasibility to recruit and randomise 50 patients across two cardiac surgical centres and secondary outcomes included the feasibility of collecting the planned perioperative data and clinically relevant outcomes and assessments of effective patient identification, screening and recruitment.

**Results:** All 50 patients were recruited within 11 months in two centres allowing for a 13-month hiatus in recruitment due to the COVID-19 pandemic. Overall, 50/108 (46%) of eligible patients were recruited. One patient withdrew before surgery and one patient did not undergo surgery. All but one completed in-hospital and 30-day follow-up.

**Conclusions:** It is feasible to recruit and randomise higher-risk patients undergoing CABG surgery to a study comparing total inhalational and propofol anaesthesia in a timely manner and with high acceptance and completion rates.

## Introduction

Coronary artery bypass graft (CABG) surgery is the revascularisation strategy of choice for patients with multi-vessel coronary artery disease. However, cardiopulmonary bypass (CPB) and myocardial revascularisation cause ischemia-reperfusion injury, leading to myocardial and other end-organ damage. Myocardial protection has been demonstrated in experimental settings and it can be triggered by ischemic preconditioning via two main intracellular signal transduction pathways, the reperfusion injury salvage kinases (RISK) and survivor-activating factor enhancement (SAFE) pathways.^1,2^ These pathways converge in the mitochondria and act upon the mitochondrial permeability transition pore (mPTP) to favor cell survival over cell death.^3,4^ Interestingly, volatile anaesthetics mimic the activation of these myocardial protective pathways while propofol might be an inhibitor.^5,6^

Potential beneficial myocardial effects of volatile anaesthetics have been compared with intravenous agents in many clinical trials and meta-analyses, indicating some benefit on patient cardiovascular outcomes, but without a definitive answer.^7–9^ Crucially, many of these studies have examined the use of volatile anaesthesia in combination with propofol infusions compared to propofol use alone. This concomitant administration of propofol with volatile anaesthetics conflicts with the demonstrable evidence that volatile agents used during CPB without additional propofol administration can reduce postoperative markers of myocardial injury, when compared with propofol use alone.^10,11^ Also, in both clinical and experimental studies, propofol has been shown to restrict myocardial protective processes.^6,12–15^

The MYRIAD study randomised 5400 patients undergoing CABG to either total intravenous anaesthesia (TIVA) or volatile anaesthesia.^16^ However, within the volatile anaesthesia group, there were high rates (59%) of coadministration of propofol during the anaesthesia maintenance. In addition, patients undergoing off-pump procedures and patients with a low risk of ischemia reperfusion injury were included in this study. The authors reported no significant difference for relevant clinical outcomes, including mortality at 1-year, between the two groups.^16^ A post-hoc analysis of the MYRIAD study, however, demonstrated a lower rate of myocardial infarction (MI) with hemodynamic instability and a reduction of 1-year cardiac mortality in patients receiving volatile anaesthetics. The authors conclude that these post-hoc results indicate potential clinically relevant cardioprotective effects by volatile anaesthetics, and they suggest that this should be further assessed, despite neutral effects on all-cause mortality.^17^

There has been recent demonstration that the administration of volatile anaesthesia during CPB is feasible, with oxygenator exhaust volatile concentrations correlating with arterial blood concentrations, and attainment of adequate hypnosis and amnesia by this technique.^18,19^

Overall, there is sufficient equipoise, even amongst noncardiac surgery, that a large randomised controlled trial (RCT) is underway for anaesthesia maintenance with volatile anaesthetic agents compared with TIVA including numerous clinical outcomes (VITAL; ISRCTN62903453). Currently there is heterogeneity in clinical practice for anaesthesia in cardiac surgical procedures in the UK and in Europe with approximately 50% of patients receiving intravenous anaesthesia alone without volatile anaesthesia.^20,21^ Therefore, demonstration of a clinically important reduction in myocardial injury with a volatile-based anaesthetic technique would have far-reaching practice implications.

We intend to assess whether a volatile-only anaesthetic strategy, i.e. total inhalational anaesthesia, for cardiac bypass surgery on CPB, compared with a propofol anaesthetic strategy, reduces postoperative cardiovascular morbidity (major adverse cardiovascular and cerebral events, MACCE) as the overarching hypothesis. We describe here the findings of a feasibility study designed to investigate recruitment and protocol adherence to the randomised treatment allocation.

## Methods

This study received Research Ethics Committee approval (London – Chelsea, 19/LO/1071, 2^nd^ August 2019) and was prospectively registered with EudraCT (No.: 2019-000171-16) and ClinicalTrials.gov (NCT04039854). All participants were aged 18 years and above and provided written informed consent.

We undertook a single-blind randomised controlled trial to assess the feasibility of a subsequent larger study, which will aim to assess whether volatile anaesthetics, as the sole hypnotic agent for general anaesthesia during elective CABG (with or without valve) surgery will reduce postoperative myocardial injury and cardiovascular morbidity in high-risk adult patients, compared with propofol anaesthesia. We sought to determine whether recruitment, protocol adherence and data collection would be feasible, as well as piloting clinically relevant outcomes for the proposed full trial.

### Patients

This feasibility study was conducted at two sites: King’s College Hospital and St Thomas’ Hospital, both London, UK.

Adult patients (aged ≥18 years) undergoing CABG surgery on CPB, with or without concomitant valvular surgery, and with an additive European System for Cardiac Operative Risk Evaluation (EuroSCORE) ≥5 were eligible to participate. Patients were excluded if they were pregnant or breastfeeding, allergic to propofol, had known sensitivity to any volatile anaesthetic agent (isoflurane, desflurane, sevoflurane, or other halogenated anaesthetic) had known or suspected malignant hyperthermia, were currently receiving any agent known to interfere with myocardial preconditioning (glibenclamide, allopurinol, theophylline or nicorandil), or were included in any other clinical trial of an investigational medicinal product within the last 3 months. Patients were allowed to be enrolled in registry or observational studies whilst participating in our study.

Eligible patients were approached preoperatively for written informed consent. Consenting patients were randomised on the morning of surgery, in 1:1 allocation by a secure web-based system (Sealed Envelope Ltd, London, UK), to the propofol (control) arm or the volatile anaesthetic (intervention) arm.

### Trial Conduct

Routine pre-anaesthetic care was the same across both arms, guided by the local evidence-based protocol. Similarly, induction of anaesthesia was based on the usual care of cardiac surgical patients with a bolus of propofol as the commonly used anaesthetic agent. Patients randomised to the volatile anaesthetic arm were assigned to receive an inhalational halogenated ether for maintenance of anaesthesia. These included isoflurane (1-chloro-2,2,2-trifluoroethyl difluoromethyl ether) or sevoflurane (fluoromethyl-2,2,2-trifluoro-1-ethyl ether), and were at the discretion of the attending anaesthesiologist. The volatile anaesthetic agent was delivered prior to and following CPB by inhalation, and during CPB through the oxygenator oxygen inflow of the CPB machine. The dose of the volatile anaesthetic agent was titrated to standard clinical end points suggesting sufficient depth of anaesthesia and a Bispectral Index (BIS) of 30-60. Volatile agent administration concluded with the end of surgery. Sedation for the transfer to and on the cardiac intensive care unit was with propofol infusion. Deviations from treatment assignment were permitted, and collected as part of routine data collection.

Patients randomised to the propofol arm received propofol for maintenance of anaesthesia, delivered via intravenous infusion, and titrated to maintain adequate depth of anaesthesia clinically, and a BIS of 30-60. As for the volatile arm, postoperative sedation in the postoperative care unit was with continued propofol infusion until tracheal extubation.

All other anaesthetic care was conducted in line with consensus-based locally approved institutional methods, including the use of other agents routinely used in cardiac anaesthesia, such as benzodiazepines, neuromuscular blocking agents, analgesics and vasoactive agents. Cardiac surgical care followed evidence-based institutional protocols, including CPB, as did postoperative management on the cardiac intensive care unit (ICU).

### Data Collection & Trial Outcomes

Preoperative patient characteristics and operative details (including surgical, perfusion and anaesthetic management) were collected by an unblinded research nurse team, whereas postoperative management and relevant clinical outcomes were collected by a different research nurse team, which was blinded to the treatment allocation. A further 30-day telephone follow-up was performed by a blinded research nurse.

The primary outcome was an assessment of the feasibility of the study protocol, assessed by:

i. determination of the likely rate of recruitment at two centres with the aim to complete recruitment within 12 months;
ii. the identification of potential recruitment barriers with the existing protocol.

Secondary outcomes included:

i. an assessment of effective patient identification, screening and recruitment;
ii. the feasibility of collecting the planned perioperative data in more than 95% of enrolled patients at the 30-day follow-up point;
iii. an assessment of trial processes, including outcome measures;
iv. an assessment of feasibility of collecting a number of clinically relevant outcomes until 30 days after surgery, including low cardiac output syndrome (LCOS), Stroke, MI or death from any cause, cardiac related mortality, postoperative atrial fibrillation (AF) requiring treatment, ICU and hospital length of stay, patients reported disability and Quality of Life (European Quality of Life – 5), (Suppl Table 1).

### Sample Size & Statistical Analysis

We aimed to recruit 50 patients between the two centres, within an estimated timeframe of 12 months. As a feasibility study, no power calculations were performed.

A Consolidated Standards of Reporting Trials (CONSORT) diagram was selected to display the key data relating to the primary outcome and several of the secondary outcomes, including recruitment, randomisation, adherence to allocation and follow-up.

Patient characteristic data is presented with descriptive statistics by allocation arm. Operative and anaesthetic management, alongside postoperative outcomes is presented similarly. Continuous variables are presented as means (with standard deviation) or medians (with interquartile range) as appropriate together with the number of observations. Categorical variables are presented as number of observations and percentages.

## Results

### Primary Outcomes

50 participants were recruited across both centres within 11 months of active recruitment, from November 2019 until November 2021, with a 13-month hiatus (March 2020 – April 2021) due to the COVID-19 pandemic. A single site was open to recruitment before pre-COVID with 19 patients randomised in 3.7 months. Both sites were open to recruitment for 6.9 months post-COVID; with 31 patients recruited). The pre-pandemic recruitment rate in the single site was 5.1 patients per month and following resumption the rate was 4.5 patients per month across the two sites. A CONSORT diagram for the flow of patients through the study is shown in Figure 1. Apart from the COVID-19 pandemic, no other systemic recruitment barriers were identified.

**Figure 1.**
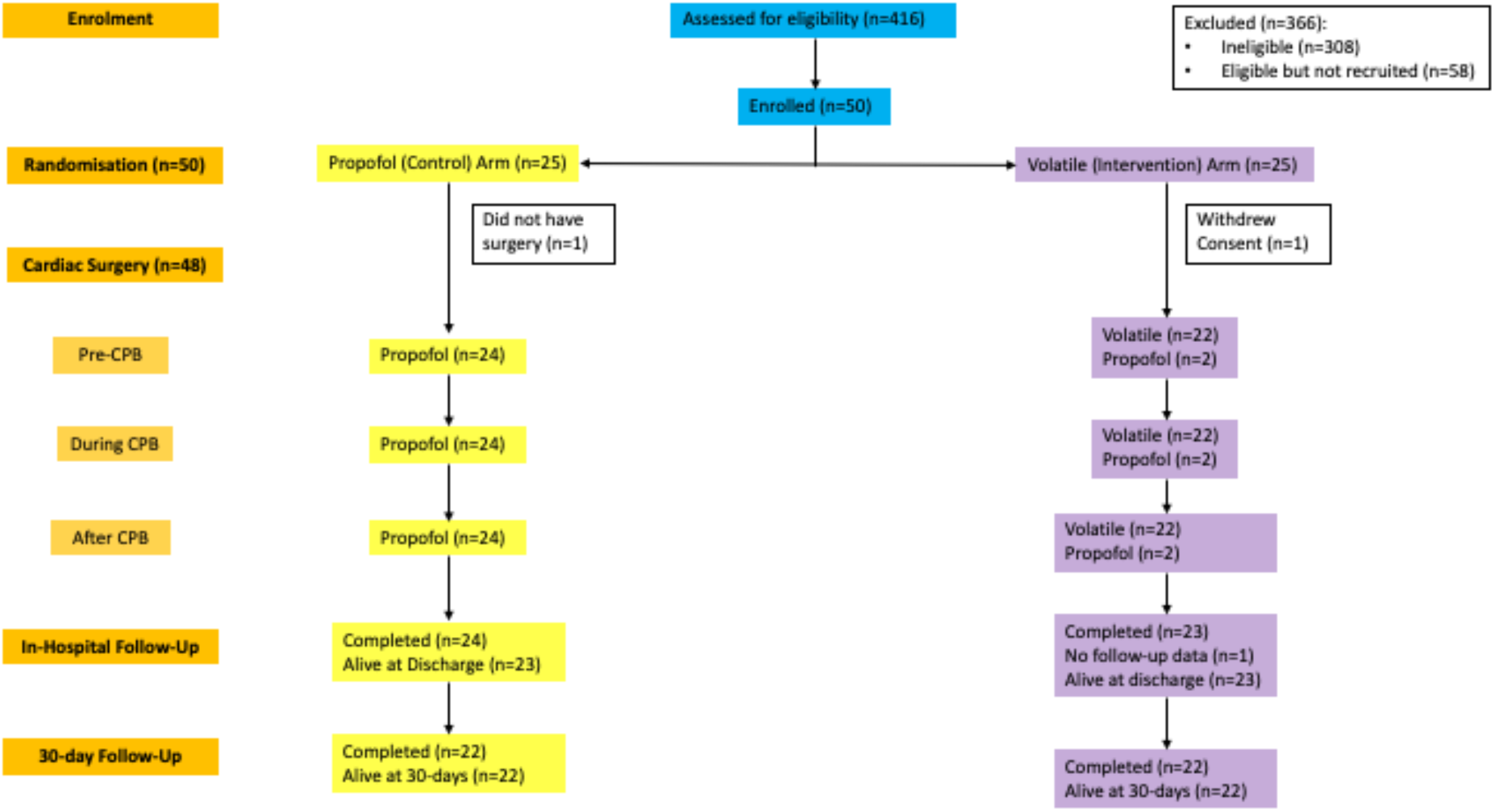
CONSORT diagram showing recruitment, randomization, adherence to treatment allocation and follow-up retention

### Secondary Outcomes

A total of 416 patients were screened during the study period. 308 (74%) were ineligible, 14% (n=58) were eligible but not recruited, and 12% (n=50) were eligible and successfully recruited. 50/108 (46%) of eligible patients were recruited to the study.

All 50 recruited patients underwent randomisation, with one withdrawing consent prior to surgery, and one patient not undergoing surgery. Of the remaining 48 patients, 47 completed in-hospital and 30-day follow-up.

Recruitment, retention and management by treatment allocation are summarised in Fig 1. In the propofol arm, all 24 patients were managed as per allocation throughout the operative period. In the volatile arm, 22/24 patients received treatment as per allocation with two protocol violations where propofol was administered.

Patients’ preoperative characteristics are summarized in Table 1. Intraoperative details and clinical outcomes are shown in Tables 2 and 3. Data completeness was good for the majority of perioperative variables including pre-and intra-operative variables as well as clinical outcomes including LCOS, AF, ICU and hospital length of stay, MACCE and cardiac-related 30-day mortality (Tables 1-3). Overall, the median time at the time point of the 30-day follow up was 33 [30-54] days in the propofol arm, and 37.5 [31-49] days in the volatile arm.

**Table 1.** Patient characteristics by treatment assignment

|  |  |  | Propofol<br>(n=25) | Volatile<br>(n=25) |
| --- | --- | --- | --- | --- |
| Age years | Mean (SD) |  | 73.0 (8.5) | 73.6 (9.1) |
|  | Missing data |  | 1 (4%) | 1 (4%) |
| Male | n (%) |  | 18 (72%) | 20 (80%) |
|  | Missing data |  | 1 (4%) | 0 |
| Ethnicity n (%) | White |  | 21 (84%) | 23 (92%) |
|  | Black |  | 1 (4%) | 0 (0%) |
|  | Asian |  | 1 (4%) | 1 (4%) |
|  | Missing data |  | 2 (8%) | 1 (4%) |
| BMI $kg/m^2$ | Mean (SD) | | 27.0 (5.8) | 28.8 (4.4) |
|  | Missing data |  | 2 (8%) | 3 (12%) |
| Preoperative BP<br>*mmHg* | Mean (SD) | Systolic | 134 (21) | 132 (19) |
|  |  | Diastolic | 72 (10) | 71 (12) |
|  | Missing data |  | 2 (8%) | 3 (12%) |
| Preoperative HR<br>*bpm* | Mean (SD) |  | 66 (9) | 64 (9) |
|  | Missing data |  | 2 (8%) | 3 (12%) |
| CCS Angina Grade n<br>(%) | 0 |  | 6 (24%) | 7 (28%) |
|  | 1 |  | 3 (12%) | 2 (8%) |
|  | 2 |  | 5 (20%) | 6 (24%) |
|  | 3 |  | 7 (28%) | 7 (28%) |
|  | 4 |  | 2 (8%) | 2 (8%) |
|  | Missing data |  | 2 (8%) | 1 (4%) |
| NYHA Stage n (%) | I |  | 6 (24%) | 8 (32%) |
|  | II |  | 7 (28%) | 10 (40%) |
|  | III |  | 10 (40%) | 4 (16%) |
|  | IV |  | 0 (0%) | 1 (4%) |
|  | Missing data |  | 2 (8%) | 2 (8%) |
| CVS Comorbidity | Arrhythmia | n (%) | 6 (24%) | 2 (8%) |
|  |  | Missing data | 2 (8%) | 2 (8%) |
|  | Hypertension | n (%) | 19 (76%) | 15 (60%) |
|  |  | Missing data | 1 (4%) | 2 (8%) |
|  | Previous MI | n (%) | 13 (52%) | 10 (40%) |
|  |  | Missing data | 1 (4%) | 1 (4%) |
| Smoking Status n (%) | Current |  | 3 (12%) | 4 (16%) |
|  | Previous |  | 12 (48%) | 10 (40%) |
|  | Never |  | 8 (32%) | 10 (40%) |
|  | Missing data |  | 2 (8%) | 1 (4%) |
| Other Comorbidity | COPD | n (%) | 1 (4%) | 3 (12%) |
|  |  | Missing data | 1 (4%) | 1 (4%) |
|  | CKD | n (%) | 6 (24%) | 3 (12%) |
|  |  | Missing data | 1 (4%) | 1 (4%) |
|  | DM n (%) | None | 14 (56%) | 19 (76%) |
|  |  | Diet-controlled | 1 (4%) | 0 (0%) |
|  |  | Oral Medication | 4 (16%) | 3 (12%) |
|  |  | Insulin | 5 (20%) | 2 (8%) |
|  |  | Missing data | 1 (4%) | 1 (4%) |
|  | TIA | n (%) | 4 (16%) | 5 (20%) |
|  |  | Missing data | 1 (4%) | 1 (4%) |
| Previous Surgery | n (%) |  | 1 (4%) | 1 (4%) |
|  | Missing data |  | 1 (4%) | 2 (8%) |
| Preoperative Medications n (%) | Aspirin |  | 16 (64%) | 18 (72%) |
|  | P2Y12 Antagonist |  | 4 (16%) | 6 (24%) |
|  | Beta-blocker |  | 16 (64%) | 15 (60%) |
|  | CCB |  | 9 (36%) | 7 (28%) |
|  | ACEi |  | 11 (44%) | 12 (48%) |
|  | ARB |  | 5 (20%) | 4 (16%) |
|  | Diuretic |  | 2 (8%) | 5 (20%) |
|  | Anticoagulant |  | 11 (44%) | 4 (16%) |
|  | Metformin |  | 6 (24%) | 5 (20%) |
|  | Sulfonylurea |  | 3 (12%) | 0 (0%) |
|  | Insulin |  | 5 (20%) | 2 (8%) |
|  | Missing data |  | 1 (4%) | 1 (4%) |
| Preoperative Laboratory Results | CBG *mmol/L* | Mean (SD) | 6.0 (1.2) | 6.7 (1.9) |
|  |  | Missing data | 12 (48%) | 9 (36%) |
|  | Creatinine *micromol/L* | Mean (SD) | 96 (26) | 107 (63) |
|  |  | Missing data | 1 (4%) | 1 (4%) |
| | Platelets $\times 10^9/L$ | Mean (SD) | 232 (61) | 211 (52) |
|  |  | Missing data | 1 (4%) | 1 (4%) |
|  | LVEF % | Median [IQR] | 52 [45-56] | 49 [40-55] |
|  |  | Missing data | 7 (28%) | 7 (28%) |
| Preoperative Cardiac Parameters (Imaging & ECG) | AF | n (%) | 1 (4.2%) | 3 (12.5%) |
|  |  | Missing data | 1 (4%) | 2 (8%) |
|  | Median [IQR] | 16 [14-17] | 15.5 [14-20] |  |
|  | Missing data | 0 (0%) | 1 (4%) |  |
| WHODAS Score | Median [IQR] |  | 16 [14-17] | 15.5 [14-20] |
|  | Missing data |  | 0 (0%) | 1 (4%) |
| EQ-5D – Health Status (0-100) | Median [IQR] |  | 72 [50-78] | 75 [65-82] |
|  | Missing data |  | 1 (4%) | 0 (0%) |

*ACEi – Angiotensin-Converting Enzyme Inhibitor; AF – Atrial Fibrillation; ARB – Angiotensin-Receptor Blocker; BMI – Body Mass Index; BP – Blood Pressure; CBG – Capillary Blood Glucose; CCB – Calcium Channel Blocker; CCS – Canadian Cardiovascular Society; CKD – Chronic Kidney Disease; COPD – Chronic Obstructive Pulmonary Disease; CVS – Cardiovascular System; DM – Diabetes Mellitus; ECG – Electrocardiogram; EQ-5D – EuroQOL-5D; HR – Heart Rate; IQR – Interquartile Range; LVEF – Left Ventricular Ejection Fraction; MI – Myocardial Infarction; NYHA – New York Heart Association; TIA – Transient Ischaemic Attack; WHODAS – World Health Organisation Disability Assessment Schedule*

**Table 2.**
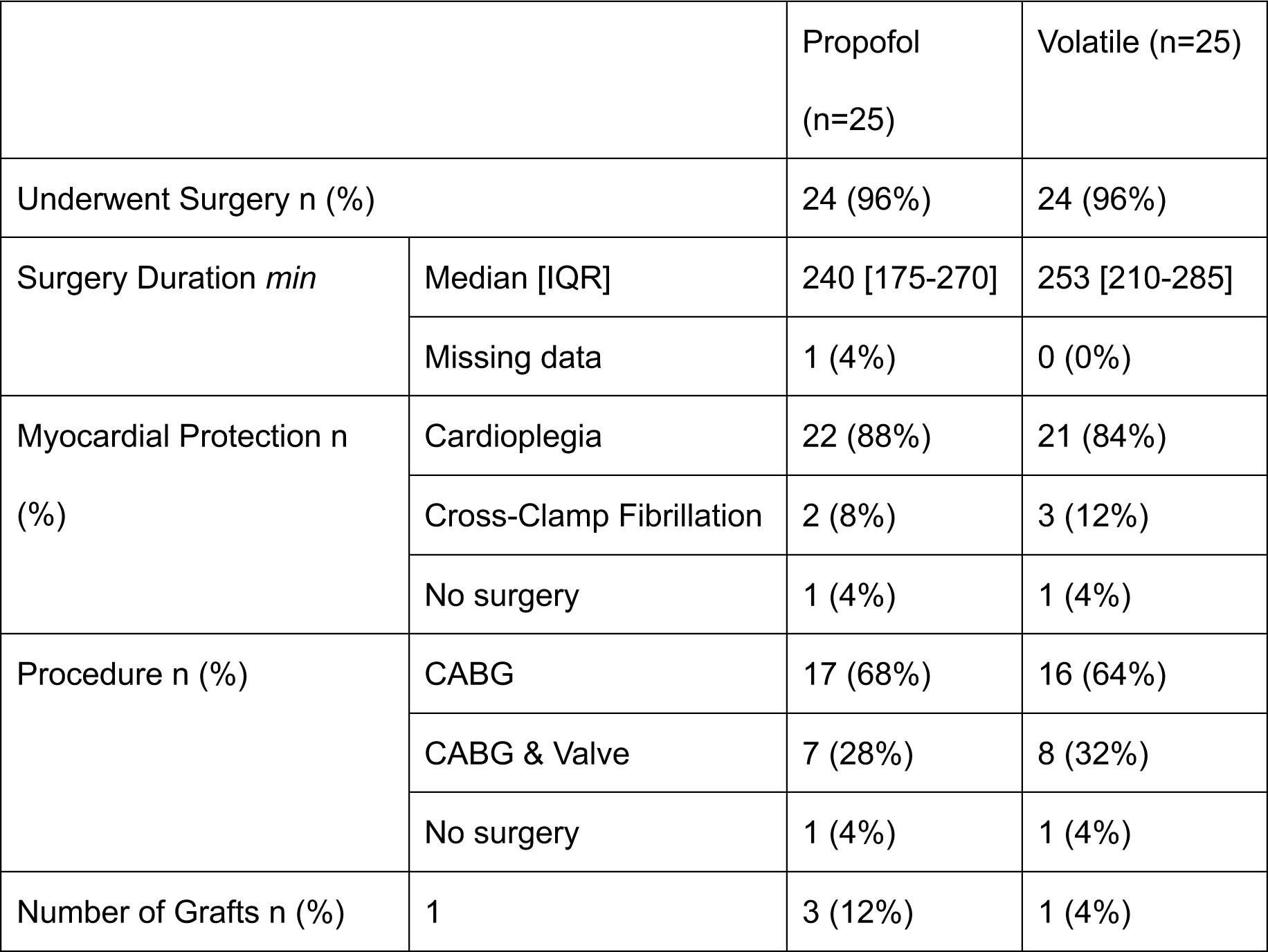
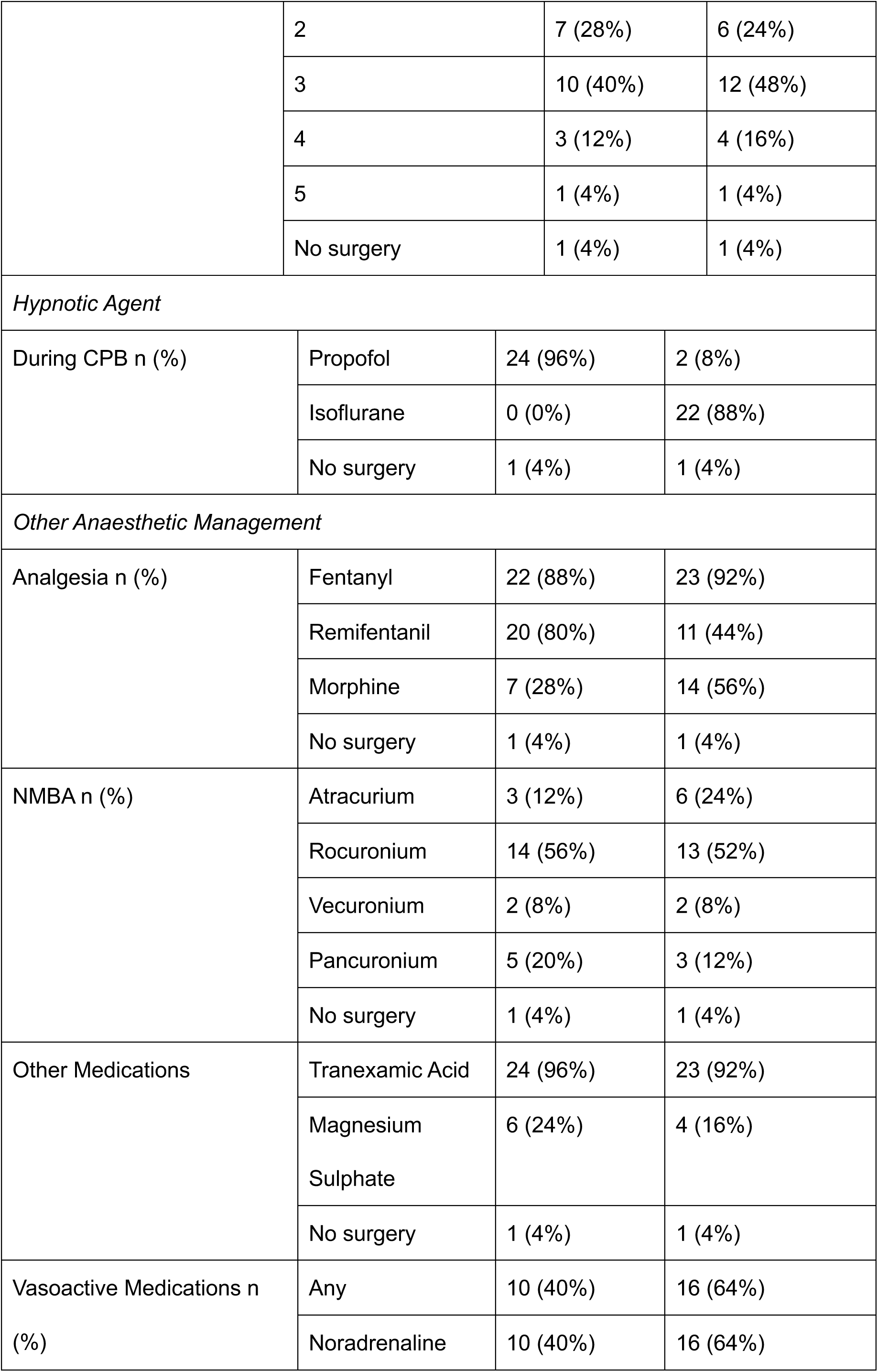
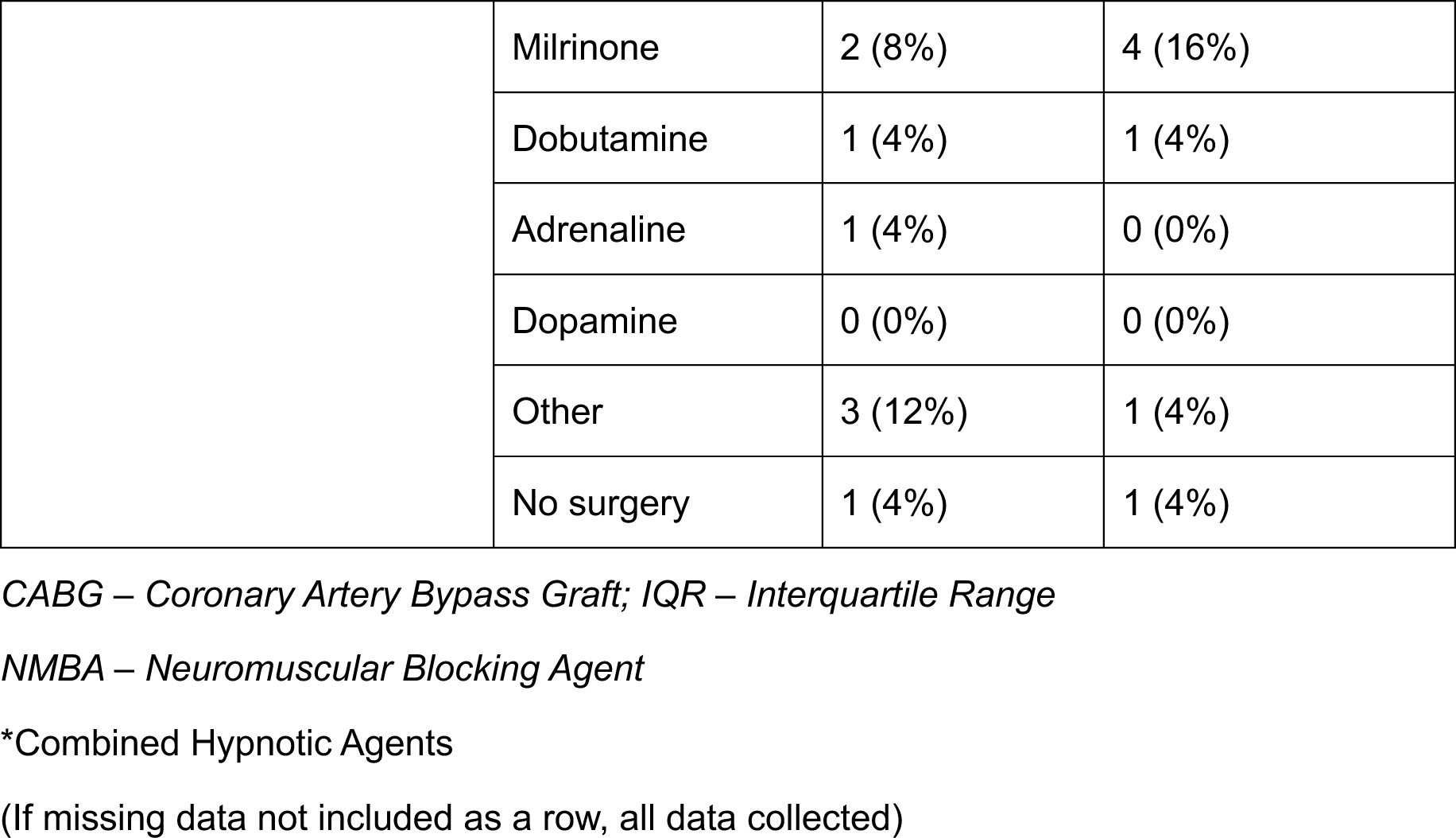
Intraoperative surgical and anaesthetic details

**Table 3.** Clinically Relevant Outcomes by Treatment Assignment

|  |  | Propofol Arm (n=25) | Volatile Arm (n=25) |
| --- | --- | --- | --- |
| LCOS n (%) | Present | 4 (16%) | 4 (16%) |
|  | IABP Criteria | 0 (0%) | 1 (4%) |
|  | Vasoactive Criteria | 4 (16%) | 3 (12%) |
|  | Missing data | 0 (0%) | 1 (4%) |
| AF n (%) | Present | 2 (8%) | 5 (20%) |
|  | Missing data | 0 (0%) | 1 (4%) |
| ICU LOS | Days Median [IQR] | 2 [2-3] | 2 [2-4] |
|  | Missing data | 0 (0%) | 1 (4%) |
| Hospital LOS | Days Median [IQR] | 7 [5-11] | 10 [7-14] |
|  | Missing data | 0 (0%) | 1 (4%) |
| Myocardial Injury | Troponin T (ng/L) |  |  |
|  | Elevated TnT (>100x 99<sup>th</sup> URL) | 4 (16%) | 4 (16%) |
|  | Preoperative TnT Median [IQR] | 19 [11-36] | 25 [14-41] |
|  | Preop Missing data | 2 (8%) | 1 (4%) |
|  | 6hr TnT Median [IQR] | 936 [632-1189] | 777 [550-1087] |
|  | 6hr TnT Missing data | 2 (8%) | 2 (8%) |
|  | Day 1 TnT Median [IQR] | 578 [389-821] | 504 [304-757] |
|  | Day 1 TnT Missing data | 2 (8%) | 1 (4%) |
|  | Day 2 TnT Median [IQR] | 326 [232-453] | 331 [202-589] |
|  | Day 2 TnT Missing data | 3 (12%) | 2 (8%) |
|  | Completed 4 TnT samples | 18 (78.3%) | 19 (79.2%) |
| MACCE n (%) | Present | 7 (28%) | 4 (16%) |
|  | Stroke | 0 (0%) | 1 (4%) |
|  | MI | 5 (20%) | 4 (16%) |
|  | Death | 2 (8%) | 1 (4%) |
|  | Missing data | 0 (0%) | 1 (4%) |
| Cardiac-related mortality at 30-days n (%) | Present | 1 (4%) | 1 (4%) |
|  | Not recorded | 0 (0%) | 1 (4%) |
| WHODAS | Recorded at 30-days n (%) (Surviving patients, n=22 in each group) | 19 (76%) | 20 (80%) |
|  | Median [IQR] | 16 [14-21] | 20 [15-24] |
|  | Change Median [IQR] | 2.0 [-2.0 – 6.0] | 2.5 [-1.0 – 9.0] |
|  | Missing data | 3 (12%) | 2 (8%) |
| EQ-5D-5L | Recorded at 30-days n (%) (from surviving patients, n=22 in each group) | 19 (76%) | 20 (80%) |
|  | Missing data | 3 (12%) | 2 (8%) |
| EQ-5D-5L Health Status | Median [IQR] | 75 [65-85] | 75 [60-80] |
|  | Change in Median [IQR] | 4.0 [-5.0 – 23.0] | -2.5 [-10.0 – 7.5] |
| Days alive and at home until 30-days after surgery* | Median [IQR] | 24 [21-30] | 22.5 [18-30] |
|  | Missing data | 0 (0%) | 1 (4%) |

\*Of surviving postoperative patients in each group (n=22)
*AF – Atrial Fibrillation; AKI – Acute Kidney Injury; EQ-5D -5L – EuroQOL-5D-5L; ICU – Intensive Care Unit; IQR – Interquartile Range; LCOS – Low Cardiac Output Syndrome; LOS – Length of Stay; MACCE - Major Adverse Cardiac and Cerebrovascular Events; MyC – Myosin-binding protein C; POD – Postoperative delirium; PPC – Postoperative pulmonary complications; URL – Upper Reference Limit; WHODAS – World Health Organisation Disability Assessment Schedule*

## Discussion

The primary outcome for this study was the feasibility of adequate patient recruitment comparing propofol with total inhalational anaesthesia for cardiac bypass surgery. Fifty patients were recruited across an 11-month period, with an enforced hiatus due to the COVID-19 pandemic, and thus feasibility of recruitment was confirmed. The initial rate of recruitment, prior to the pandemic, was 5.1 patients per month at a single center followed by a rate of 4.5 patients after the pandemic across the two sites. Based upon the actual recruitment rate in this study with 50 patients recruited in two centers within one year, a recruitment in excess of 1000 patients across 20 UK sites would be achievable in 36 months. This is bearing in mind that the two pilot centers in this feasibility trial should be considered as high recruiting centers, and an overall lower recruitment number was therefore assumed in this conservative assessment. The recruitment time could be reduced with an increased number of centers participating, e.g. to 2.5 years with 25 participating centres.

One of the secondary outcomes was an assessment of the screening and participant identification processes. These proved to be effective, and of the eligible patients identified, 46% were recruited. This compares similarly with 47% in the ERICCA study and less favorably with the MYRIAD trial (77.5%).^16,20^ The inclusion criteria in the ERICCA study and MYRIAD trial would explain the difference in recruitment rates, as the ERICCA study only included higher-risk patients with a EuroSCORE ≥ 5, which is the same cut-off as in this feasibility study, whereas the MYRIAD trial included elective CABG patients irrespective of preoperative risk. Of note, these were two large RCTs, rather than feasibility studies.

Of those participants that were randomised and underwent surgery (n=48), 47 (98%) patients completed follow-up at 30-days. This figure is within the 95% target for completing follow-up. Furthermore, assessment of the patient characteristics (Table 1), reveals a recognizable cohort of cardiac surgical patients with typical clinical co-morbidities.

We also sought to assess the feasibility of our trial processes. In the propofol arm, all patients undergoing surgery were managed throughout the operative period as per treatment allocation. In the volatile arm, 22/24 (92%) patients randomised to the volatile arm were managed as per treatment allocation that underwent surgery. These results appear to be suggestive of a high degree of feasibility for the management protocol, given the absolute values of protocol adherence, and the relative comparison with other studies. The MYRIAD RCT was a pragmatic comparison of volatile anaesthetics with TIVA for intraoperative anaesthesia management in patients undergoing elective CABG, to assess impact on mortality at 1-year. In the volatile anaesthesia group, 98% of patients received volatile agents, but, in addition, 59% of participants received intravenous hypnotics for maintenance. In the TIVA group, 99% of patients received per protocol maintenance of anaesthesia, with 3% receiving volatile anaesthetic agents.^16^ Based upon the level of treatment concordance demonstrated in this pilot study with 92% of patients receiving volatile anaesthetics only for their maintenance (compared to only 41% in the MYRIAD trial), the protocol is feasible for a larger study.

For the proposed clinically relevant outcomes, there was good data completeness for the majority of variables, such as LCOS, AF, length of stay, MACCE, cardiac-related mortality and days alive and at home at 30-days. The primary outcome for our proposed larger study will be MACCE at 12 months, and therefore high rates of complete data for many of these outcomes is vital.

Concerning cardiac biomarker levels, a similar median value of Troponin T at 24h postoperatively was seen in the propofol arm of this study, when compared with a previous study of sevoflurane versus TIVA for on-pump elective CABG surgery, primarily looking at length of stay.^11^ However, there was a much lower median level of Troponin T in the volatile arm in that study, which, in contrast to our study, included low-risk patients.^11^

Further comparison of myocardial injury values across studies is particularly difficult given disparity in the study interventions (the idiosyncrasies of the anaesthesia regimens), as well as the specifics of the study population, surgical techniques, and other facets of management. However, other clinically defined outcomes may be more readily compared, despite the small size of our study. When compared to our study, the MYRIAD study had a lower incidence of cardiac-related death at 30-days (0.7% and 0.9% in the volatile and TIVA arms respectively, compared with 4% in both arms of our study) and in a slightly more restrictive composite outcome of non-fatal MI and death at 30-days (5.0% and 4.7% in the volatile and TIVA arms, compared with the MACCE criteria (stroke, MI and death) in our study, with an incidence of up to 28%).^16^ This may indicate that our inclusion criteria may select for a higher risk of postoperative cardiovascular injury and may therefore enable the detection of a significant difference between the arms, where previous studies have failed to do so.^7,22–23^

Our results reveal that there was an incidence of LCOS, a clinical state indicating myocardial injury, in 4/25 patients (16%), and this is an agreement with the previously described incidence of LCOS (13.5%) after CABG.^24^

Anaesthetic agents, and in particular nitrous oxide (N_2_O), play an important role regarding environmental sustainability, which has been comprehensively reviewed.^25^ However, it was demonstrated that volatile halogenated anaesthetics, such as isoflurane or sevoflurane, make only a minute contribution to greenhouse gas radiative forcing (0.01-0.02% of the radiative effect that results from increases in CO_2_ by human activity), which is in contrast to N_2_O.^26^ Therefore, moving away from inhalational anaesthesia (with either isoflurane or sevoflurane) to TIVA may negatively affect the long-lived carbon to the atmosphere because of the vast quantity of plastic needed for TIVA.^26^ Consequently, there are no strong environmental motivated reasons in favor of the usage of either agent, TIVA or volatiles (i.e. isoflurane or sevoflurane).

Regarding a future large RCT comparing propofol versus total inhalational anaesthesia, a possible approach to the combination of both, clinical outcomes and biochemical markers of myocardial injury and infarction, would be the use of a composite endpoint with a win-ratio approach, and our pilot data will be very useful to inform this decision.^27^

The question whether volatile anaesthetics protect the myocardium with relevant clinical outcomes in higher risk patients undergoing on-pump CABG surgery should be addressed in a large RCT.^28^ We therefore conducted this study, assessing feasibility and demonstrating that it is indeed feasible to recruit elective cardiac surgery patients to a randomised study examining markers of myocardial injury and clinical cardiac outcome variables between those assigned to an intravenous anaesthetic regimen versus a total inhalational anaesthesia. The reported rates of recruitment, adherence to anaesthetic management by group assignment and completion of follow-up have demonstrated that a large scale RCT within a reasonable timeframe is possible with our protocol.

## Data Availability

All data produced in the present study are available upon reasonable request to the authors

## Acknowledgments

This study was supported by the National Institute of Academic Anaesthesia (WKR0-2018-0025).

---

**Suppl Table 1.**
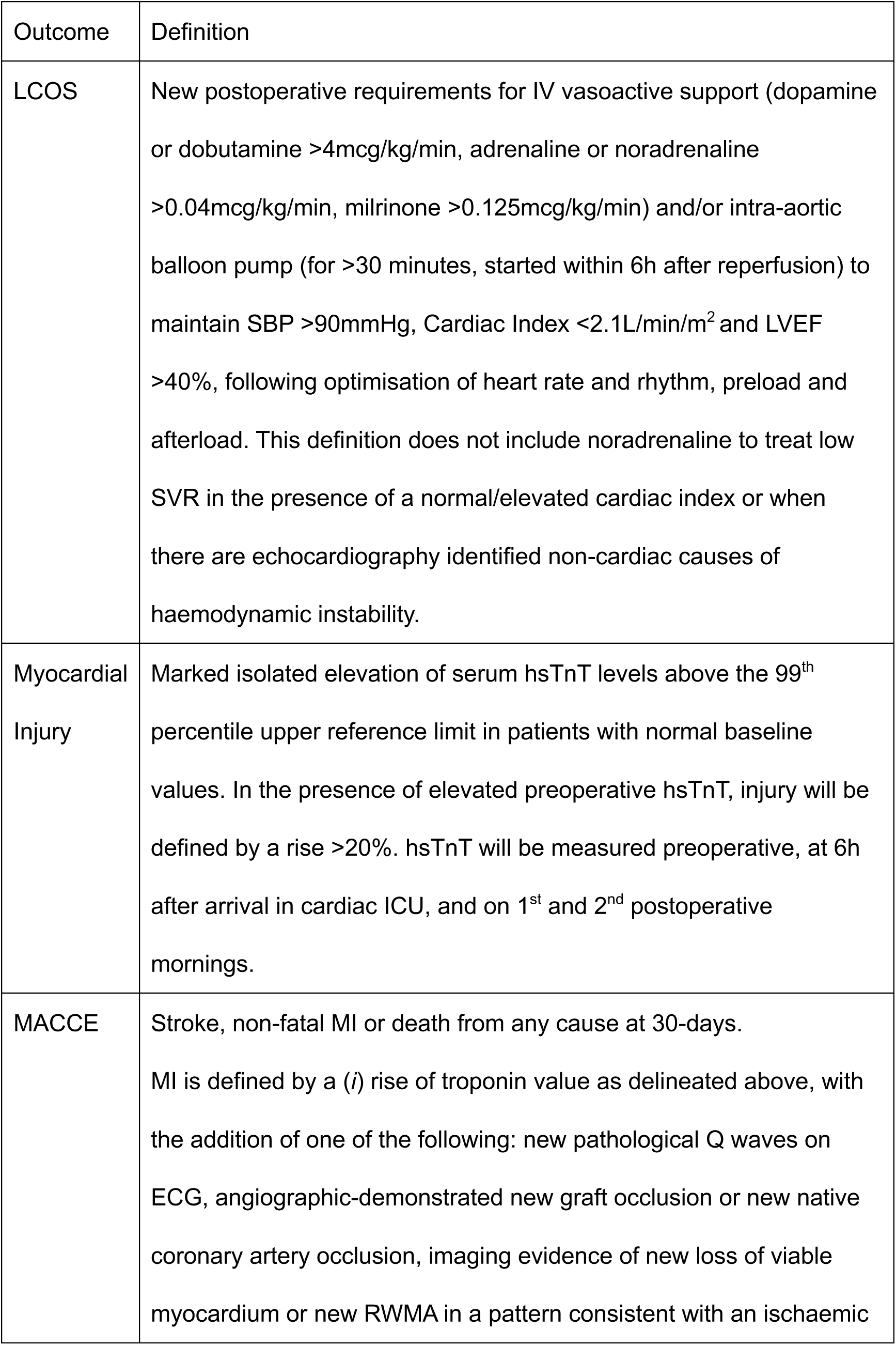
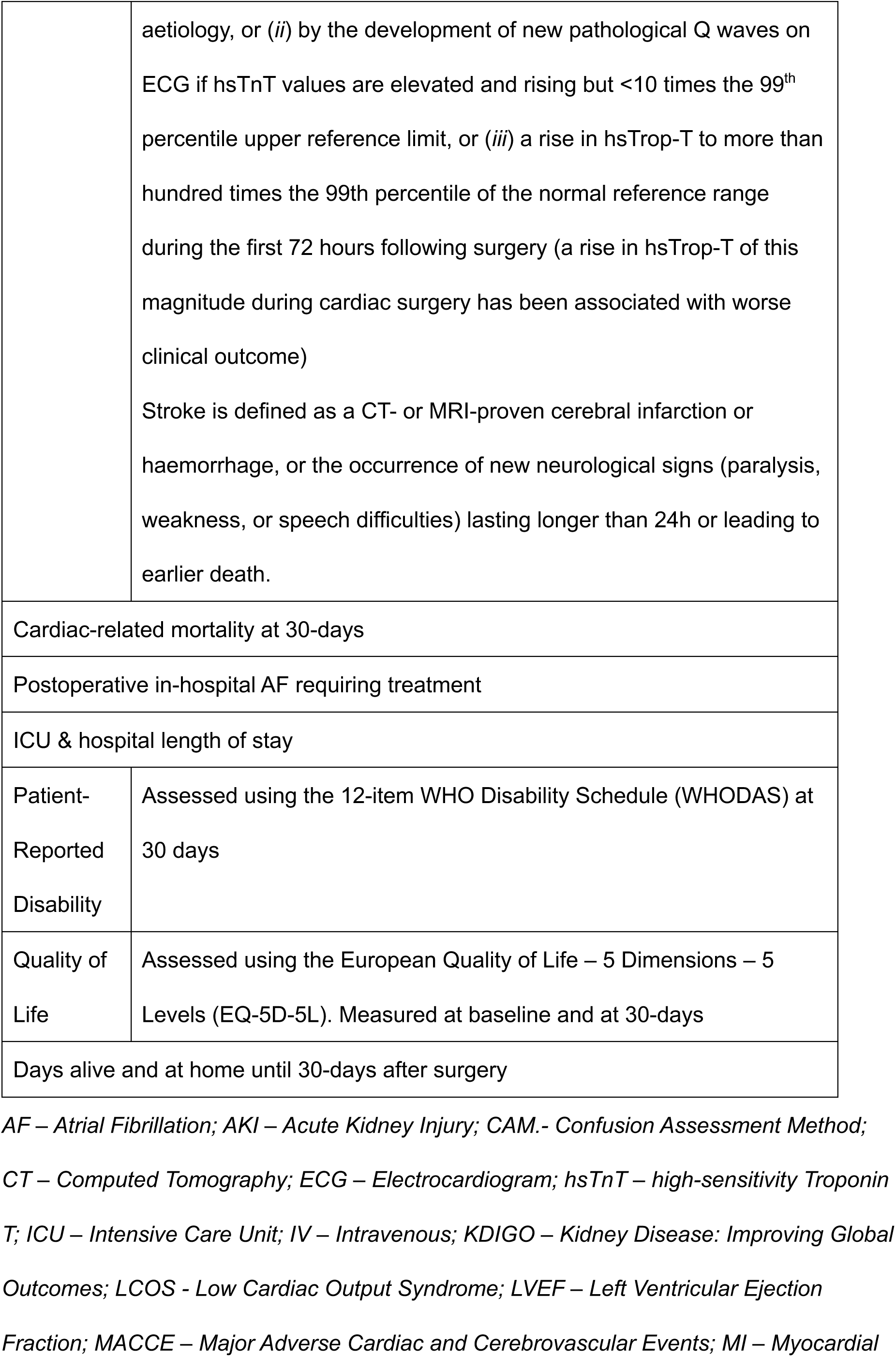
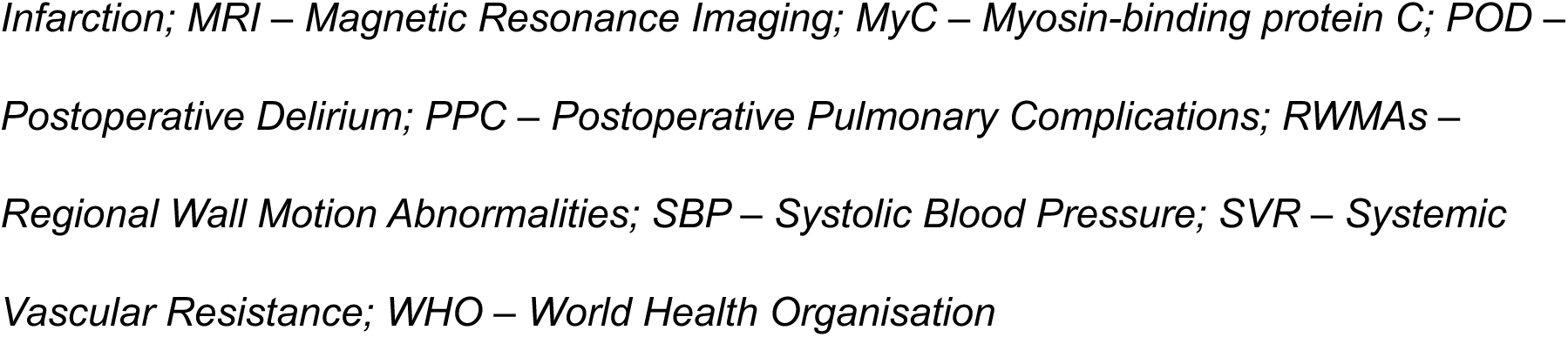
Clinically relevant outcomes constituting secondary outcomes, alongside definitions

